# Hydrocephalic Brain Volume Estimation from Low-Field MRI: Topologically-Enriched Cross-Modal Enhancement and Segmentation

**DOI:** 10.64898/2026.08.17.26360618

**Authors:** Srijit Mukherjee, Kelsey Templeton, Steven J. Schiff, Vishal Monga

## Abstract

**Objective:** Accurate volumetric analysis of brain and cerebrospinal fluid (CSF) is essential for monitoring hydrocephalus, a significant pediatric neurological condition. While computed tomography (CT) provides high-quality volumetric assessment, the associated ionizing radiation poses risks, especially for children. Low-field magnetic resonance imaging (LF-MRI) offers a safer, more accessible alternative, particularly in resource-constrained settings. However, its lower resolution and an increased likelihood for structural distortions complicate accurate segmentation. This study aims to demonstrate that reliable volumetric measurements can be obtained from LF-MRI, comparable to CT, enabling safer and more frequent monitoring of hydrocephalic infants.

**Approach:** We propose **EnSegNet-Cross**, a cross-modality guided enhancement-aware segmentation network for brain volume analysis using LF-MRI. The framework relies on high-fidelity CT data during training but requires only LF-MRI at inference. At the heart of this innovation, lies a novel cross modal topological penalty to minimize discrepancies between predicted LF-MRI and CT structures. A central contribution is the integration of a 3D topological loss based on persistent homology, which penalizes topological discrepancies in CSF regions (specifically CSF holes formed by enclosed brain parenchyma) between CT and LF-MRI segmentations. This embedding of structural priors facilitates generalization across heterogeneous clinical cases while obviating the need for CT data at inference time, leading to more anatomically coherent and topologically faithful segmentations.

**Main Results:** On a curated cohort of hydrocephalic infants with paired LF-MRI and CT scans, including infectious and non-infectious causes, EnSegNet-Cross consistently outperformed state-of-the-art machine learning alternatives by achieving the highest Dice Score (0.8532 ± 0.03) and Volume Score (0.9318 ± 0.03), and robustly handled challenging cases with confounding factors (Dice 0.8340 ± 0.03, Volume 0.9111 ± 0.05). Leveraging CT-derived topological priors allowed EnSegNet-Cross to succeed in anatomically complex scenarios where conventional models fail.

**Significance:** EnSegNet-Cross offers a reliable, interpretable solution for brain–CSF segmentation, as demonstrated in complex hydrocephalus cases. This study demonstrates that high-fidelity volumetric estimates can be achieved using only LF-MRI, facilitating frequent, radiation-free monitoring. By bridging the fidelity gap between low-quality LF-MRI and high-resolution CT through clinically grounded enhancement and topological supervision, EnSegNet-Cross provides a robust clinical tool for brain volumetric analysis in hydrocephalus infants using LF-MRI.

## Introduction

Hydrocephalus is a neurological disorder characterized by the accumulation of cerebrospinal fluid (CSF) within the brain, resulting in elevated intracranial pressure, progressive head enlargement in infants, and potential long-term neurocognitive deficits [1–3]. Although this condition poses a significant challenge in pediatric neurosurgical care globally, the condition is particularly prevalent in low-resource settings, with Sub-Saharan Africa reporting approximately 180,000 new infant cases annually, predominantly due to postinfectious etiologies [2, 4]. If left untreated, the excess fluid can lead to permanent brain damage and developmental impairments. Standard treatment includes ventriculoperitoneal shunt (VPS) placement or endoscopic third ventriculostomy (ETV) with or without choroid plexus cauterization (ETV+CPC). Recent clinical trials [5] have demonstrated that brain volume recovery post-treatment is strongly correlated with improved neurodevelopmental outcomes, underscoring the clinical importance of accurate brain volumetric assessment. This is achieved by assessing deviations from normative pediatric brain volume growth curves, derived from healthy infants’ datasets, which provide a critical reference for evaluating treatment efficacy and monitoring brain development trajectories in hydrocephalic infants [6–8] Quantitative neuroimaging, particularly segmentation of MRI and computed tomography (CT) scans into brain tissue and CSF compartments, offers a viable pathway for such evaluations. Therefore, reliable, automated segmentation frameworks tailored to abnormal hydrocephalic brain anatomy are essential for advancing diagnostic, prognostic, and interventional strategies, particularly in resource-constrained environments.

CT is commonly employed for diagnostic brain imaging due to its widespread availability and high spatial resolution, making it well-suited for volumetric analysis [4, 9]. However, its reliance on ionizing radiation poses significant risks for infants and young children, including increased susceptibility to radiation-induced brain tumors [10, 11]. In contrast, low-field MRI (LF-MRI) systems, such as the portable Hyperfine scanner, provide a safer and more accessible alternative, particularly in resource-limited settings [12, 13]. Recent studies [14–16] have demonstrated that even low-resolution brain imaging can effectively support the management of infant hydrocephalus. Despite their advantages, LF-MRI often suffer from reduced resolution and signal quality, complicating tasks such as classification and segmentation.

Hydrocephalus is marked by substantially enlarged and distorted ventricles compressing the surrounding brain parenchyma [17]. These structural alterations introduce significant segmentation challenges in LF-MRI, whose inherent low resolution and low signal-to-noise ratio (SNR) further obscure tissue boundaries. The difficulty is compounded in postinfectious hydrocephalus (PIH), which accounts for over 50% of pediatric hydrocephalus cases in sub-Saharan Africa [2, 3]. PIH presents a constellation of complex imaging features, including intraventricular debris, calcifications (readily resolved with CT), abscesses, and loculated CSF compartments, that severely confound conventional segmentation pipelines [2, 18].

Our study was motivated by a clinically realistic and technically challenging scenario: the availability of paired LF-MRI and high-resolution CT scans during training, but only LF-MRI during real-world inference. This training was enabled by a curated dataset of hydrocephalic patients‡, where CT labels serve as anatomical ground truth for volumetric calculation. Our goal was the development of a cross-modality learning solution capable of learning from high-fidelity CT data during training while generalizing to LF-MRI inputs alone during inference and deployment. Addressing this modality gap requires a model that not only captures the pathological complexity of hydrocephalus but also encodes transferable representations from CT into the LF-MRI domain. Figure 1 illustrates the cross-modal learning problem addressed in this work. We propose the AI framework **EnSegNet-Cross**, which performs CT-supervised enhancement of low-field MRI (LF-MRI) images, followed by segmentation during training. At inference time, the model enables accurate brain and CSF segmentation, along with volumetric estimation, using only low-resolution LF-MRI inputs.

**Figure 1.**
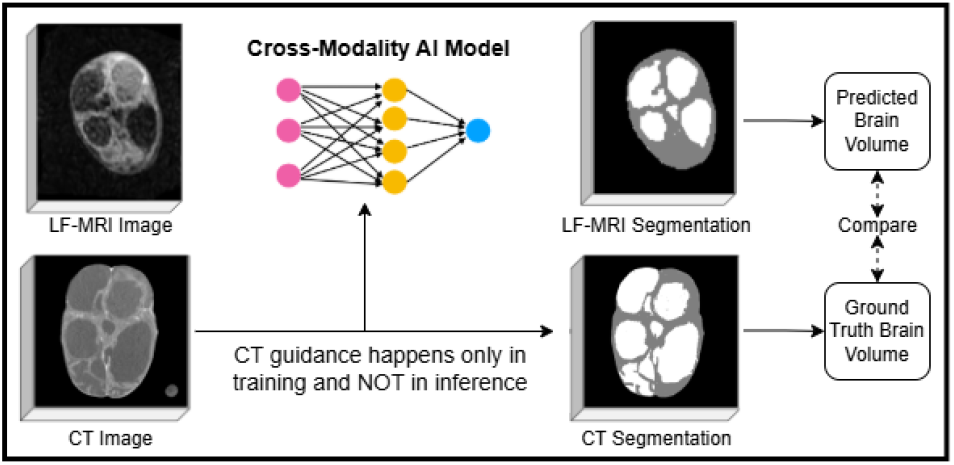
The Cross-Modality Learning Problem

### Related Works

The field of automated medical image segmentation has evolved significantly, from early rule-based computer-aided diagnosis (CAD) systems employing classical image processing and statistical learning techniques [20–24], to deep learning models capable of learning hierarchical features directly from high-dimensional data. Segmentation in medical imaging has also been proven to be important for volumetric analysis and medical image classification tasks [25] such as infection diagnosis in hydrocephalic infants [26, 27]. Convolutional Neural Networks (CNNs) [19, 28– 32] and Vision Transformers [33–36] now constitute the state-of-the-art in segmentation across diverse clinical tasks. More recently, foundation models such as the Segment Anything Model (SAM) [37–39] have enabled prompt-driven zero-shot segmentation, inspiring a new class of approaches [40–44] that encode domain knowledge through visual prompts. However, these advances typically presume clean, high-quality imaging data, limiting their applicability leading to poorer performance (Figure 2C) in real-world, low-resource environments such as portable LF-MRI, where low resolution, and disease-induced spurious signals (Figure 2B) are prevalent compared to their corresponding high-quality counterparts such as CT (Figure 2A - 1st row). This limitation is especially pronounced in hydrocephalus, where segmentation on LF-MRI is underexplored and often requires manual annotation [45, 46]. Prior work in LF-MRI enhancement has focused on global denoising or signal amplification [47–51], generally ignoring disease-specific anatomical distortions. Cross-modality learning offers an appealing alternative by allowing high-fidelity modalities such as CT to guide segmentation on lower-quality counterparts.

**Figure 2.**
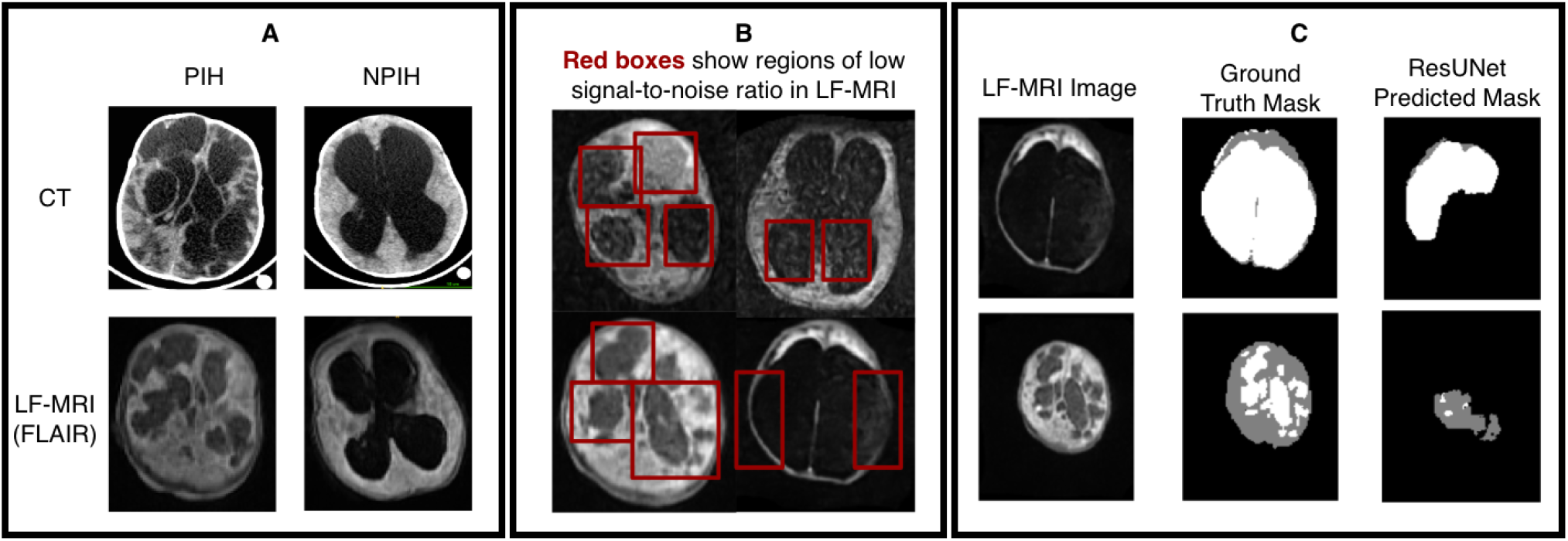
(A) Paired dataset comprising LF-MRI (FLAIR) and corresponding CT scans. (B) Red boxes exhibit regions of image quality degradation due to disease-related spurious signals (debris within CSF), and low resolution in LF-MRI scans. These factors obscure boundaries between CSF, brain tissue, and skull, making accurate segmentation challenging. CSF loculations distort global brain geometry, further complicating segmentation. (C) State-of-the-art models such as ResUNet[19] struggle to segment accurately for hydrocephalus in LF-MRI specific conditions. This underscores the need for a novel neural architecture for this problem.

### Motivation

Most of the cross-modal learning methods [52–61] largely rely on unsupervised domain adaptation or symmetric style transfer between high-fidelity modalities (e.g., T1 and T2 MRI, CT and PETCT), assuming high image quality, clean anatomical correspondence, and shared information. They are designed for environments where both source and target modalities are of high fidelity, in which segmentation supervision is available only in the source domain, while the target domain remains unlabeled. These strategies falter in LF-MRI of hydrocephalic patients, where image degradation, anatomical deformation, and CSF hyperintensities undermine alignment and correspondence, as evident in Figure 2B. Our approach fundamentally departs from existing works in two crucial ways: 1) We have ground truth segmentation labels for LF-MRI, in contrast to the works above, where the target domain (like LF-MRI) does not have access to the ground truth labels. Consequently, our model is not trained under a source-labeled/target-unlabeled paradigm. 2) CT and LF-MRI are not peers in fidelity or completeness. CT is not a domain to adapt from, but a structural reference that compensates for quality imperfections in LF-MRI. We therefore reconceptualize cross-modality not as domain adaptation but as anatomical augmentation, using CT exclusively during training to statistically enhance LFMRI and impose topological correctness. Rather than translating style or appearance, CT serves as a source of anatomical priors, guiding LF-MRI enhancement through structural regularization. Our training objective combines supervised segmentation loss on LF-MRI with CT-informed regularizers that promote anatomical plausibility via topological and structural constraints. We introduce a novel topological loss that leverages global topology of CSF regions, particularly enclosed CSF cavities formed by compressed brain parenchyma, to align predicted LF-MRI segmentations with high-fidelity CT references. This hybrid loss enforces both volumetric agreement with ground truth CT and consistency of brain–CSF boundaries.

### Our Contribution

We introduce **EnSegNet** and **EnSegNet-Cross**, two modular architectures for robust brain–CSF segmentation in hydrocephalus infants using LF-MRI. Both models are built upon an enhancement-driven prompting paradigm that conditions enhancement through hydrocephalus etiology-guided anatomical priors, followed by segmentation with the Segment Anything Model (SAM) [37] with disease-informed unsupervised prompt selection. EnSegNet is a fully LF-MRI-based segmentation framework, while EnSegNet-Cross incorporates cross-modal supervision during training by leveraging high-quality CT data (only during training). This design allows EnSegNet-Cross to transfer geometrical and structural knowledge from CT into LF-MRI representations using distribution alignment and topology-aware loss functions. Specifically, the main contributions are:

#### 1) Disease-Aware Enhancement Guided Unsupervised Prompt Generation

We propose an Enhance Module that suppresses debris-related hyperintensities inside CSF pockets and increases brain–CSF contrast in LFMRI, producing spurious-signal-reduced images with FLAIR-like signal characteristics. These enhanced images are then provided to a Prompt Module built upon the pre-trained Segment Anything Model (SAM) [37], where disease-informed and modality-aware unsupervised prompts guide high-quality segmentation. The coupling of enhancement and prompt selection enables robust segmentation by leveraging pathology-specific and imaging-derived priors in our frameworks.

#### 2) Topologically Enriched CT-Guided Cross-Modal Segmentation

In EnSegNet-Cross, we introduce a Cross Module that uses CT images only during training to improve LF-MRI representation learning. Within this framework, a Distribution Matching Module (DMM) aligns CT features with the LF-MRI domain to provide effective cross-modal supervision, while geometric and topological consistency is imposed through a CT-guided topological loss derived from anatomical structures (during training only). Together, these components result in structurally faithful LF-MRI segmentation for volumetric hydrocephalus quantification.

#### 3) Experimental Validation and Clinical Insights

We evaluate EnSegNet and EnSegNet-Cross on a curated cohort of hydrocephalic infants with paired LF-MRI and CT scans, spanning both infectious and non-infectious etiologies. Across Dice overlap and volume correlation metrics, both EnSegNet and EnSegNet-Cross outperform the state-of-the-art alternatives.

## Methods

### A. Ethics Statement

This project was performed on deidentified human image data following informed consent under the oversight of the Institutional Review Boards of Yale University, Penn State University, and the Research Ethics Committee of the Mbarara University of Science and Technology. The CT scans were deidentified before sharing with researchers. LF-MRI scans were processed in two data streams – one with full personal health information directed to the local server at the point-of-care for clinical use, and another fully deidentified stream to the Hyperfine Cloud, where researchers were able to access these data under agreement with Hyperfine and with IRB approval. For infant studies, IRB approval was also obtained to share birth dates and dates of scans to characterize images at young ages.

### B. Dataset and Data Preprocessing

The dataset for this study was collected at CURE Children’s Hospital of Uganda (CCHU), a nationwide neurosurgical referral center located in Mbale, in the eastern region of Uganda. It is the same dataset of pathological hydrocephalus images as previously used in [26]. The dataset comprises 47 infants with paired low-field fluid-attenuated inversion recovery (FLAIR) MRI and computed tomography (CT) images: 28 were labeled as postinfectious hydrocephalus (PIH), and 19 were labeled as non-postinfectious hydrocephalus (NPIH). Each infant’s FLAIR sequence contains approximately 40 images, while there are, on average 100 images for each CT scan sequence. The classification of PIH and NPIH was based on established clinical criteria [18]. For both the FLAIR and CT images, we also have ground truth segmentation masks annotated by medical professionals, which are divided into three classes: brain tissue, cerebrospinal fluid (CSF), and the rest as separate regions. All the scanned images were collected in DICOM format. Brain region (including brain tissue and CSF) was extracted for FLAIR and CT using state-of-the-art skull stripping methods [27].

### B. Notation

Each infant subject is associated with a sequence of 2D CT slices and 2D LF-MRI slices, accompanied by manually annotated ground-truth segmentation maps provided by expert clinicians. Let *X*_lf_ ∈ **R**^*H*×*W*^ denote a preprocessed LF-MRI slice and *X*_ct_ ∈ **R**^*H*×*W*^ denote a preprocessed CT slice from the same subject, where *H* and *W* represent the image height and width, respectively. The associated pixel-wise segmentation masks are denoted as *M*_lf_ and *M*_ct_, where *M*_lf_, *M*_ct_ ∈ {0, 1, 2}^*H*×*W*^ encodes three semantic regions: brain tissue (label 1), cerebrospinal fluid (CSF, label 2), and non-brain/background (label 0); region-specific trinary ground-truth masks are denoted as 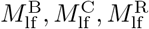 and 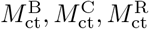 for the brain, CSF, and rest of the brain respectively; the corresponding predicted masks are given by 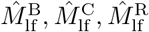 and 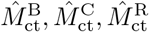. The LF-MRI input *X*_lf_ is passed through a neural network *f* (·; *θ*), parameterized by *θ*, yielding a predicted segmentation 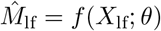. The model is optimized by minimizing a composite loss ℒ (*θ*) that includes segmentation loss terms, an enhancement loss designed to suppress pathological spurious signals, and a topological loss to enforce structural consistency of neuroanatomical features between CT and LF-MRI. We propose two variants of this network architecture: EnSegNet and EnSegNet-Cross. In EnSegNet, training is performed solely on LF-MRI slices. In contrast, EnSegNet-Cross incorporates both LF-MRI and CT slices within a cross-modality training framework, enabling the network to learn shared representations guided by anatomical priors from CT data. In both cases, the final model outputs 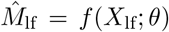, using the optimized parameters 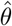, is capable of performing segmentation on LF-MRI slices independently, without requiring CT input during inference.

### D. Rationale behind our approach

LF-MRI offers a promising pathway toward democratized neuroimaging access, yet its clinical utility remains hindered by fundamental limitations in image fidelity [13]. Such difficulties are problematic in anatomically and pathologically critical regions such as CSF spaces affected by structural deformation, altered compartmental geometry, and disease-induced debris. In hydrocephalic patients, debris within CSF, expected to appear hyperintense in FLAIR sequences, often manifests as anomalous hyperintensities, compounded by LF-MRI’s inherently low resolution, which blurs tissue boundaries and leads to intensity overlaps between CSF, and brain parenchyma. These pathological distortions give rise to topologically inconsistent features, including spurious cavities, disconnected components, and ambiguous tissue interfaces, which mislead both convolutional and transformer-based state-of-the-art segmentation models that mistake noise for a structural signal. To address this core neural engineering challenge, we propose a two-tiered architecture as illustrated in Figure 3: EnSegNet, which targets modality-aware enhancement and segmentation, and EnSegNet-Cross, which leverages CT-based cross-modal supervision to impose topological integrity. EnSegNet consists of two synergistic modules: (1) a CNN-based Enhancement Module that learns to produce artifact-suppressed pseudo-FLAIR images by rendering CSF regions dark, guided by ground truth masks, and (2) a Prompt Module built on the Segment Anything Model (SAM) [37] generates anatomically grounded segmentations through intelligent prompt selection informed by hydrocephalic priors and the enhancement output. However, due to the irreducible signal-to-noise limitations of LF-MRI, enhancement and prompting alone are insufficient. EnSegNet-Cross introduces a cross-modality training paradigm wherein CT scans distribution-matched to LF-MRI via histogram specification are processed through a shared-weight encoder-decoder network, enabling co-learning and alignment of anatomical structures from both modalities. A central contribution is the integration of a 3D topological loss based on persistent homology, which penalizes topological discrepancies in CSF regions, specifically enclosed CSF cavities formed by compressed brain parenchyma between CT and LF-MRI segmentations. This embedding of structural priors facilitates generalization across heterogeneous clinical cases while obviating the need for CT data at inference time.

**Figure 3.**
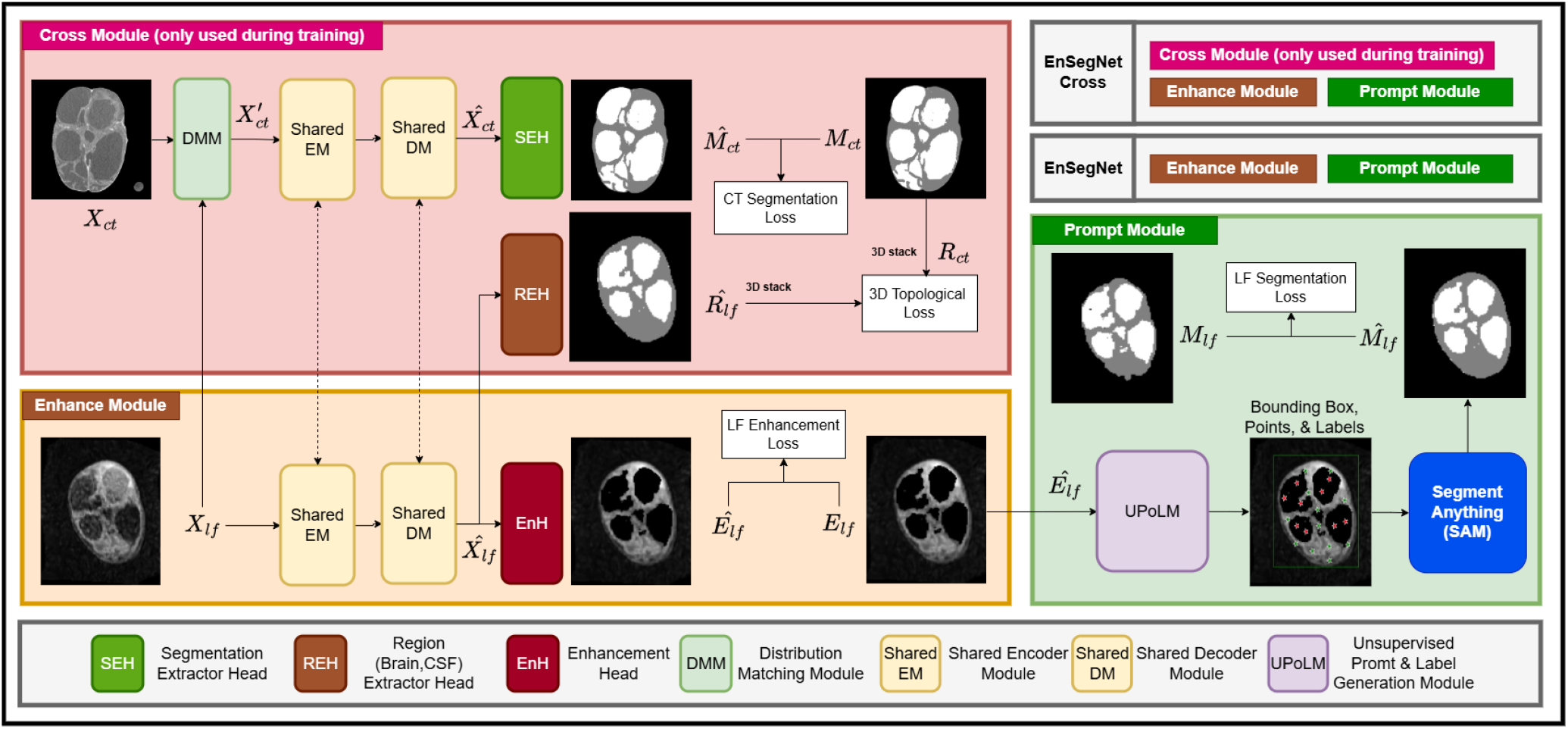
A multi-module framework for LF-MRI image enhancement and segmentation, integrating cross-module training and unsupervised prompt generation. Symbols: *X*_*lf*_ : Low-Field image, 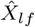: LF feature representation, *E*_*lf*_ : LF enhanced image, *Ê*_*lf*_ : LF predicted enhanced image, 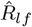: LF predicted 3D volume, *R*_*ct*_: CT ground truth 3D volume, 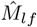: LF predicted mask, *M*_*lf*_ : LF ground-truth mask, *X*_*ct*_: CT image, 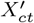: CT distribution-matched image, 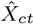: CT feature representation, 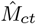: CT predicted mask, *M*_*ct*_: CT ground-truth mask. Note that EnSegNet-Cross uses CT during training only, and only uses LF-MRI during inference.

### E. EnSegNet Architecture

EnSegNet (Figure 3) introduces a two-stage architecture for artifact-robust segmentation of LF-MRI images. The input slice *X*_lf_ is first processed by a learnable *Enhance Module, f* (· | *θ*_*E*_), which suppresses pathology-induced spurious signals like debris-related hyperintensities in CSF regions, while preserving salient anatomical boundaries. The resulting enhanced image *Ê*_lf_ is then passed to a pretrained *Prompt Module, g*(· | *θ*_*P*_), based on the Segment Anything Model (SAM) [37], which generates refined prompts to guide segmentation toward clinically relevant structures, producing the final prediction 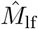. This design integrates modality-driven LF-MRI enhancement with disease-informed prompt selection within a foundation model framework, enabling robust segmentation in low SNR settings. Particularly, it preserves ventricular topology under hydrocephalic deformation, where state-of-the-art approaches often fail (Figure 2 C). We next describe the individual roles of the *Enhance Module* and *Prompt Module*.

#### 1) Enhance Module

The Enhance Module is designed to learn a structurally consistent representation of LF-MRI while suppressing pathology-induced spurious signals, particularly hyperintensities localized within cerebrospinal fluid (CSF) compartments. To this end, it employs a U-Net-based encoder–decoder architecture [28] with residual skip connections, enabling multiscale feature extraction while preserving fine-grained anatomical structure. Each LF-MRI slice *X*_lf_ is processed by the network *f* (· | *θ*_*E*_) to produce intermediate feature representations 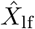, which enhance tissue contrast and maintain spatial fidelity. Region-specific activations are denoted as 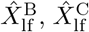, and 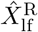, corresponding respectively to brain, CSF, and background structures. These features are passed to a dedicated enhancement head (EnH), which predicts a CSF likelihood map 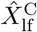. The final enhanced output is computed as 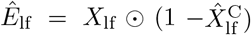, where ⊙ denotes element-wise multiplication, effectively suppressing CSF-related hyperintensities while preserving surrounding brain parenchyma. This operation emulates the signal suppression behavior of Fluid-Attenuated Inversion Recovery (FLAIR) imaging. Supervision is provided via a ground-truth enhanced image defined as 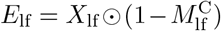, where 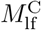 is the manually annotated CSF mask. The module is trained using a composite loss (Section Methods G) combining structural similarity and intensity alignment to ensure anatomically consistent enhancement.

#### 2) Prompt Module

The Prompt Module converts enhanced LF-MRI inputs into anatomically precise segmentation masks using the pre-trained Segment Anything Model (SAM) [37], without supervised fine-tuning. At its core is the *Unsupervised Prompt and Label Generation Module* (UPoLM), which performs disease-informed prompt synthesis directly from the enhanced output *Ê*_lf_ produced by the Enhance Module. UPoLM provides a pathology-aware, unsupervised prompting strategy that adapts to structural variability and imaging degradation in LF-MRI, particularly in hydrocephalus. The Prompt Module, defined as *g*(· | *θ*_*P*_) with SAM pre-trained weights *θ*_*P*_, takes *Ê*_lf_ as input and produces segmentation predictions 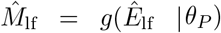 by steering SAM’s transformer-based attention through a hierarchical prompt-generation pipeline as shown in Figure 4 (B, C). As illustrated in Figure 4 (B), UPoLM first extracts a bounding box covering intracranial contents by selecting the *α*-th percentile of high-intensity voxels in *Ê*_lf_ and padding it by ±*k* pixels in each direction (*α* = 90, *k* = 20, empirically optimal for skull extraction). This bounding box is passed to SAM to generate an initial binary skull mask, which is subsequently refined through morphological hole filling to ensure a contiguous intracranial region. The enhanced image along with the skull mask (Figure 4 C) is then processed using maskSLIC (masked Simple Linear Iterative Clustering, SLIC) [62, 63], a superpixel-based segmentation algorithm specifically designed to partition irregular masked regions into spatially coherent and locally homogeneous superpixels. A superpixel is a compact cluster of neighboring pixels that share similar image characteristics, such as intensity and spatial continuity, thereby providing a structured regional representation of the image rather than treating each pixel independently. This representation is particularly advantageous in our prompt-generation framework, as it organizes the intracranial region into meaningful local compartments that facilitate reliable prompt selection. Rather than selecting prompts from random individual pixels, each superpixel serves as a candidate anatomical local region for the representative prompt point. Within our framework, each generated superpixel inside the intracranial mask is summarized by its mean intensity and subsequently clustered using *K*-means [64] to assign semantic labels. Superpixels belonging to high-intensity clusters, typically corresponding to brain parenchyma, are designated as *positive points* (green), whereas superpixels belonging to low-intensity clusters, typically corresponding to CSF are designated as *negative points* (red). The centroids of these labeled superpixels provide spatially distributed and anatomically representative point prompts. These structured positive and negative point prompts along with the skull mask, and bounding box prompts are then supplied to SAM to guide its attention toward clinically relevant brain–CSF boundaries. Following SAM inference, we obtain a brain segmentation mask corresponding to the intracranial brain parenchyma. To isolate CSF, we then use a simple anatomical decomposition strategy: the CSF region is obtained by subtracting the predicted brain mask from the skull mask. Importantly, SAM does not restrict predictions to prompted locations: prompts act as semantic anchors that condition globally structured attention, where the SAM’s Vision Transformer (ViT) image encoder applies self-attention across all image patches and the mask decoder uses cross-attention between prompt tokens and patch tokens, enabling segmentation to propagate to spatially distinct but appearance-consistent regions of the same anatomical structure, even in unprompted areas. The effectiveness of this propagation critically depends on two complementary factors: 1) Enhancement, which suppresses spurious signals and improves feature consistency across LF-MRI, providing SAM with reliable visual cues for global attention; and 2) Disease-informed prompt selection, which ensures that the prompts accurately represent anatomically and pathologically relevant regions. Together, these mechanisms enable SAM to leverage global attention for accurate brain/CSF segmentation from sparse unsupervised disease-aware prompting strategy, yielding anatomically grounded robust segmentation masks even with low SNR LF-MRI with quality degradation and variability.

**Figure 4.**
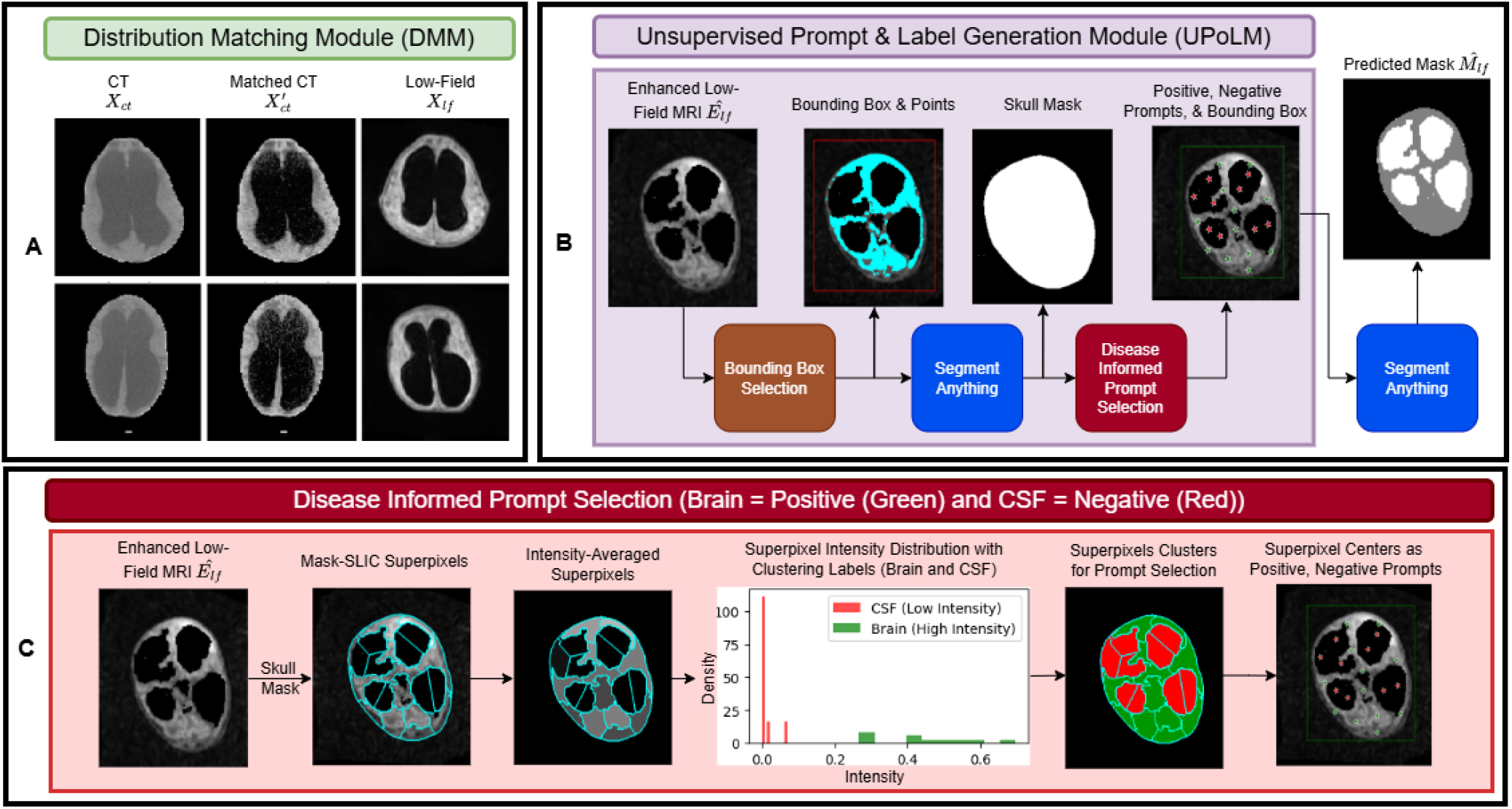
(A) Distribution Matching Module (DMM): Aligns intensity distributions of CT (*X*_ct_) to low-field MRI (*X*_lf_) using histogram matching, producing a modality-aligned representation 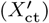 that enables shared encoder learning across modalities. (B) Unsupervised Prompt & Label Generation Module (UPoLM): Generates disease-informed prompts for the Segment Anything Model (SAM) using an initial skull mask derived from enhanced LF-MRI. Intensity-based bounding boxes define the region of interest, while superpixel clustering produces spatially coherent positive (brain) and negative (CSF) point prompts. These prompts, together with the bounding box, are provided to SAM to obtain the final segmentation mask. (C) Disease-Informed Prompt Selection: The enhanced LF-MRI within the intracranial skull region is partitioned into superpixels using MaskSLIC. Intensity-aware clustering separates high intensity and low intensity regions, from which representative superpixel centroids are selected as positive (brain) and negative (CSF) prompts. This ensures anatomically consistent guidance for robust brain–CSF segmentation during inference.

### F. EnSegNet-Cross Architecture

While EnSegNet operates on single-modality LF-MRI data, EnSegNet-Cross (Figure 3) extends this framework by incorporating computed tomography (CT) guidance during training as an auxiliary supervisory signal. This cross-modality design leverages the superior anatomical fidelity of CT, particularly its enhanced bone-tissue and tissue-fluid contrast and sharper boundary delineation to improve the structural consistency and segmentation quality of LF-MRI outputs. Crucially, CT data is used *only* during the training phase, never at inference, ensuring that the final pipeline remains entirely CT-free at deployment. To facilitate this transfer of structural priors from CT to LF-MRI, EnSegNet-Cross introduces a dedicated *Cross Module* comprising two key components: (i) a distribution-matched segmentation pathway in which Distribution Matching Module (DMM) helps intensity-align CT images with LF-MRI via histogram specification, allowing them to be processed through the same encoder-decoder architecture, and (ii) a topological alignment mechanism, wherein region-specific predictions from LF-MRI are aligned with CT-derived anatomical ground truth using a cross-modal topological loss between ground truth CT CSF mask and predicted LF-MRI CSF mask. Together, these components enable EnSegNet-Cross to learn anatomically faithful representations under cross-modality supervision, leading to improved segmentation performance in structurally degraded LF-MRI contexts.

#### 1) Cross-Module Mask Prediction

To achieve modality-invariant feature alignment within EnSegNet-Cross, we implement a cross-modal transformation based on 3D histogram matching through Distribution Matching Module (DMM) (Figure 3). For each infant subject, we consider the full volumetric CT sequence and its paired LF-MRI sequence. A global histogram transformation is computed across the entire 3D CT volume using the intensity distribution of the corresponding 3D LF-MRI volume as reference. This results in a consistent distribution-matching transformation *T* : **R** → **R** applied voxel-wise across all slices of the CT sequence. Each CT slice *X*_ct_ is thus transformed into a distribution-matched version 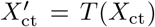 as illustrated in Figure 4 (A), where *T* (*x*) satisfies the histogram alignment criterion *C*_ct_(*T* (*x*)) ≈ *C*_lf_(*x*), for each intensity value *x* ∈ *X*_ct_. Here, *C*_ct_(·) and *C*_lf_(·) represent the empirical cumulative distribution functions (CDFs) of the full 3D CT and LF-MRI volumes, respectively. This transformation ensures global intensity consistency across modalities, mitigating domain shifts while preserving anatomical fidelity. The resulting distribution-matched CT slice 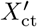 is then passed through the shared encoder-decoder network *f* (· | *θ*_*E*_), which is jointly trained on both CT and LF-MRI inputs. Finally, the encoded features 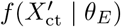 are processed through a Segmentation Extraction Head (SEH) to produce the predicted CT mask 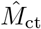. This crossmodal strategy of 3D intensity alignment facilitates shared representation learning and improves anatomical generalization across diverse imaging conditions.

#### 2) Cross-Module Topology Alignment

Following distribution matching, the CT and LF-MRI slices are processed through a shared encoder-decoder architecture *f* (· | *θ*_*E*_), enabling joint feature learning across modalities. The encoded LF-MRI feature maps, denoted 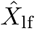, are subsequently passed through a dedicated *Region Extractor Head* designed to isolate the 3D cerebrospinal fluid (CSF) component. To enforce anatomical fidelity, we leverage the availability of paired CT-based ground truth masks 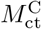 to compute a topological alignment loss. Specifically, for each infant all CSF region predictions from the LF-MRI sequence are stacked into a 3D volume 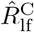, while corresponding ground-truth CT segmentations are stacked to form a 3D CSF volume 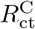. A persistent homology-based *topological loss*, denoted ℒ_topo_ (Section Methods G3), is then applied between these 3D volumes to penalize discrepancies in topological features, specifically 3D holes formed by enclosed CSF regions within brain parenchyma. This cross-modal topological supervision enforces preservation of CSF topology (fluid loculations and ventricular morphology), thereby improving the anatomical realism of LF-MRI segmentation.

### G. Loss Functions

While EnSegNet focuses exclusively on single-modality LF-MRI inputs, EnSegNet-Cross incorporates cross-modality guidance from CT scans during training to enforce anatomical fidelity and topological consistency. Importantly, EnSegNet and EnSegNet-Cross are trained under different objective functions. The loss function for EnSegNet includes only the low-field segmentation loss ℒ_LFSeg_ and the enhancement loss ℒ_Enhance_. In contrast, EnSegNet-Cross introduces two additional supervision signals: a CT segmentation loss ℒ_CTSeg_, which guides shared feature learning using CT ground-truth masks, and a cross-modality topological loss ℒ_Topo_, which aligns the 3D topology of predicted LF-MRI CSF structures with that of CT.

#### 1) Segmentation Losses

Dice loss ℒ_*Dice*_ operates on region-wise masks for both modalities. For CT:

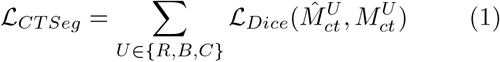

where 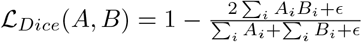 . Similarly,

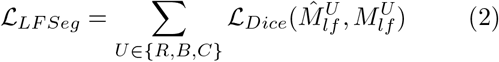

is computed for LF predictions. We use 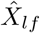 rather than 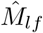 in ℒ_*LF Seg*_ to avoid propagating gradients through SAM, reducing computational cost without sacrificing accuracy. Note that 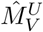, and 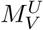 represent predicted and the ground truth segmentation masks, respectively for region *U* and imaging modality *V* as defined in Section Methods C (Notation).

#### 2) Enhancement Loss

We supervise the Enhance Module by enforcing structural similarity between the network-predicted enhanced LF image *Ê*_lf_ = *f* (*X*_lf_ | *θ*_*E*_) and the ground-truth enhanced image *E*_lf_. Concretely, we use the Structural Similarity Index (SSIM) [65], which is a well-known measure of image fidelity w.r.t a reference and normalized in the range [0, 1]. Because SSIM is to be maximized, the enhancement loss to minimize is given by

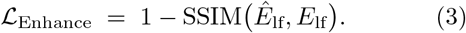

#### 3) Topological Loss (CSF) via Persistent Homology

Figures 5 and 6 motivate the use of topological loss for CSF segmentation. As shown in Figure 5, thresholding of scalar fields reveals how topological structures such as connected components and hole-like regions evolve across scales, and persistence diagrams provide a compact summary of these multiscale events. However, conventional segmentation networks often fail to preserve such global structural properties, leading to anatomically inconsistent predictions. Figure 6 illustrates how incorporating a topological loss aligns persistence diagrams between LF-MRI predictions and CT-derived ground truth masks, reducing structural discrepancies and improving segmentation consistency. This suggests that topological supervision can help enforce structural fidelity in segmentation tasks. Note that likelihood maps and masks are used interchangeably for the discussion. Prior works [66–69] have used persistent homology to extract multiscale topological features such as connected components, cycles, and voids from segmentation outputs, and compared them to ground truth using optimal transport distances between persistence diagrams. These approaches have shown improvements in tasks such as 3D cell shape analysis, brain tumor segmentation, and general medical image segmentation. In a departure from [66–68], we enforce topological consistency between distinct modalities of CT ground-truth CSF and the predicted LFMRI CSF likelihood maps. Recently, [70] suggested that the topology of ventricular surface curvature is a meaningful differentiating factor between adult normal-pressure hydrocephalus, Alzheimer’s disease, and healthy adults. To the best of our knowledge, our work constitutes the *first effort* to use topological consistency for brain tissue/CSF segmentation from a lower-quality modality such as LF-MRI. We now expand upon the pipeline shown in Figure 5, explaining in detail the mathematical formulation and intuitive reasoning behind each step. The figure provides a concrete illustration of how CSF likelihood maps are processed to yield persistence diagrams (PDs) and how these diagrams are compared via a topological loss. Our analysis operates on the image domain Ω ⊂ **R**^3^ (where CSF cavities are associated with 2D homology features), but for simplicity of explanation, we present the pipeline in the simplified setting Ω ⊂ **R**^2^ (1D homology features) in the steps below.

**Figure 5.**
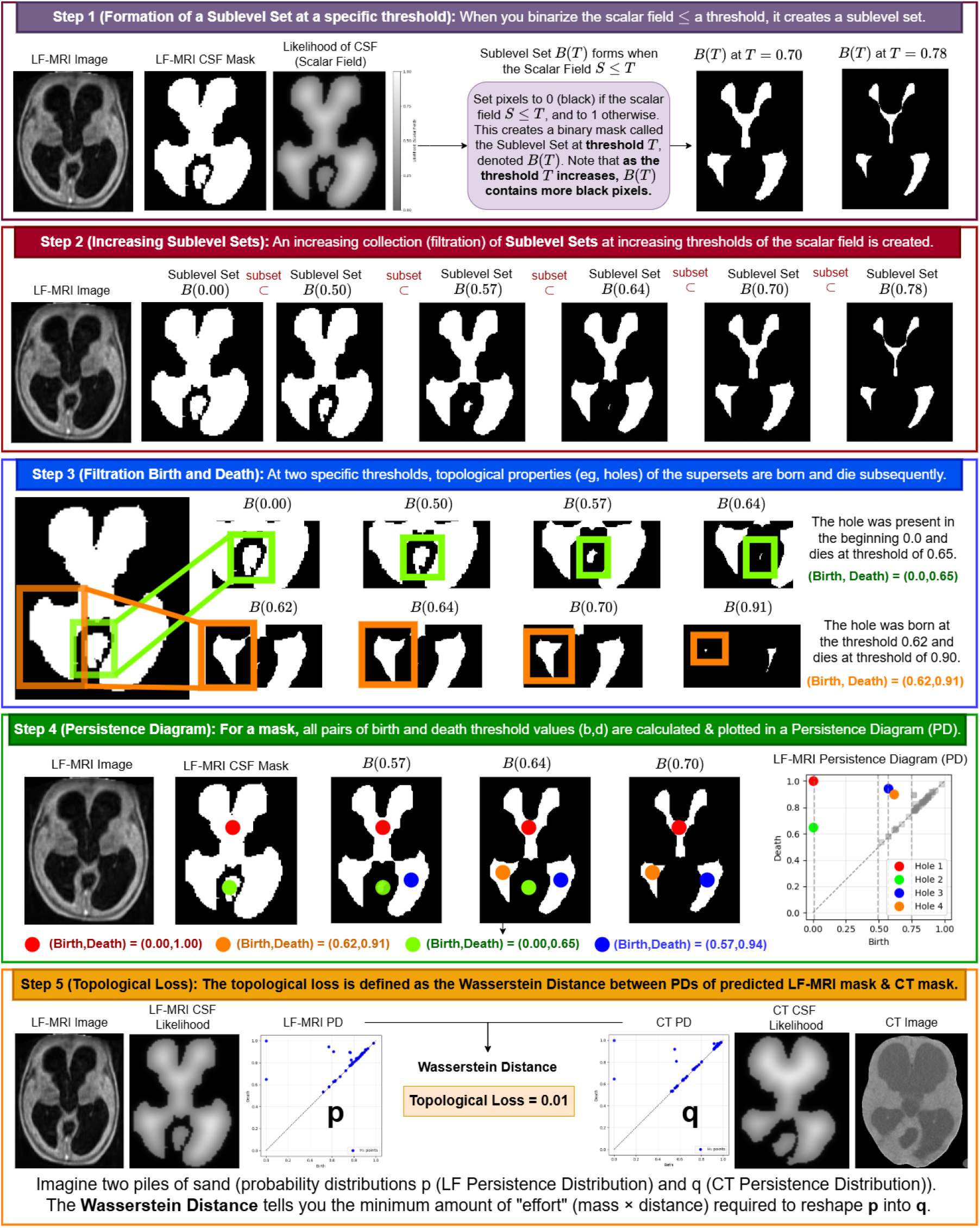
Pipeline overview. (Step 1) A scalar field is thresholded to construct sublevel sets, producing binary masks at different intensity levels that highlight regions of high (white) CSF likelihood. (Step 2) By varying the threshold, these masks form a nested increasing sequence (a filtration), where CSF regions gradually shrink and their topology evolves as CSF holes (1D holes - *H*_1_) split, and vanish. (Step 3) As the filtration progresses, topological features, specifically 1D holes (*H*_1_) are tracked. Each feature is assigned a birth *b* (when it appears) and death *d* (when it disappears) threshold values. (Step 4) These birth–death pairs (*b, d*) are summarized in a persistence diagram (PD), where each point represents a topological feature of the segmentation. The diagram provides a compact topological signature: points far from the diagonal (*b* ≈ *d*) correspond to persistent, structurally significant features, while near-diagonal points typically reflect noise. (Step 5) The topological loss is defined as the Wasserstein distance (see Figure 6) between the persistence diagram of the prediction (*p*) and that of the ground truth (*q*). Conceptually, this distance measures the minimum “effort” required to transform one diagram into the other, analogous to the work needed to reshape one distribution of mass into another (e.g., moving a pile of sand to match a target shape). In the example shown, the resulting loss is low (0.01), indicating that the predicted LF-MRI segmentation is topologically very similar to the ground-truth CT map.

**Figure 6.**
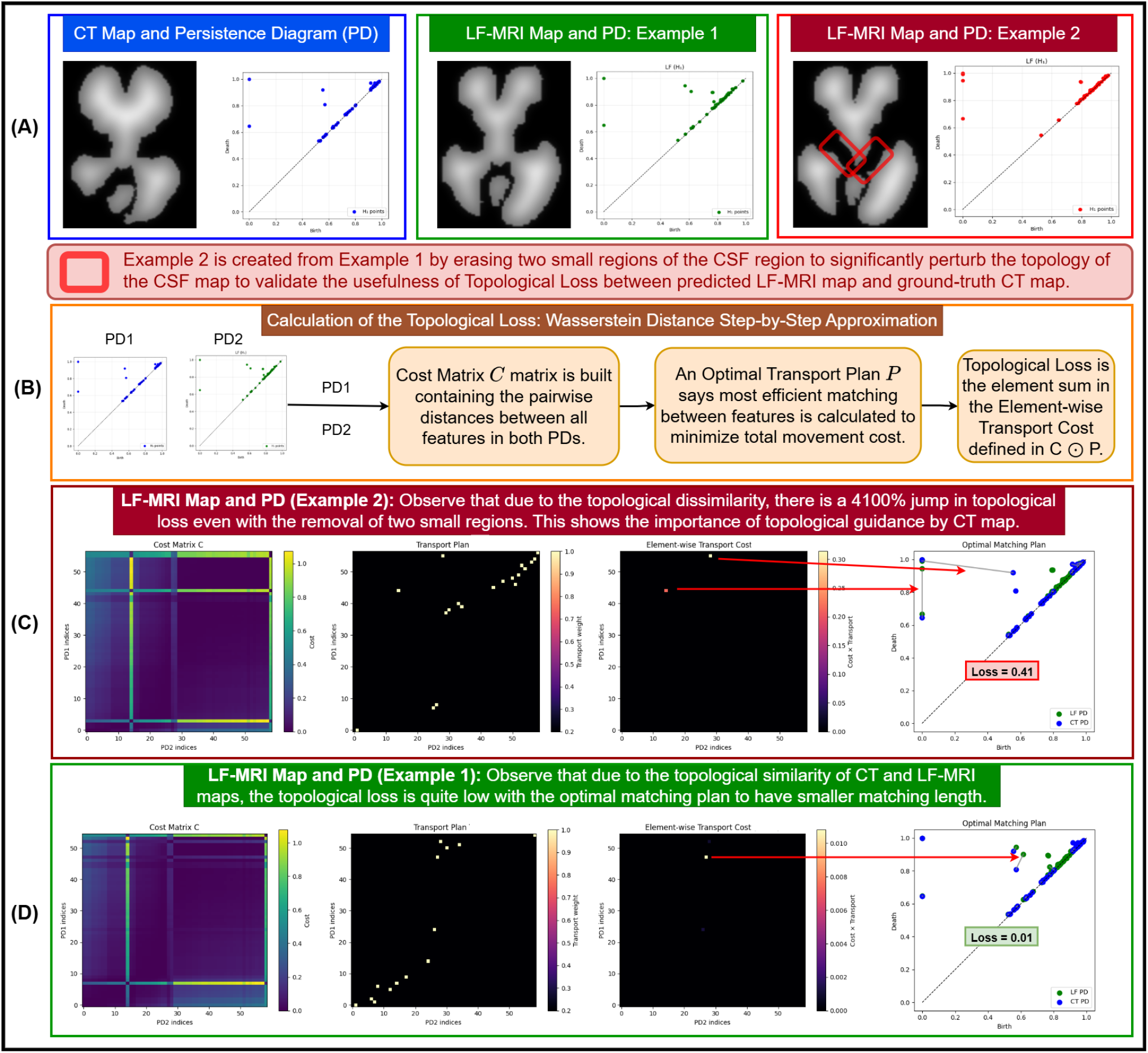
Importance and step-by-step calculation of the topological loss. (A) Persistence diagrams of three likelihood maps: a CT map (blue), a topologically faithful LF-MRI map: Example 1 (green), and Example 2 (red) constructed from Example 1 by erasing two small CSF regions to introduce spurious topological holes. (B) The topological loss equals the Wasserstein distance between PDs, computed in three steps: (i) build a cost matrix *C* whose entry (*i, j*) encodes the distance between feature *i* of the LF-MRI PD and feature *j* of the CT PD (as shown in the plots in first column of C,D below); (ii) solve for an optimal transport plan *P* (second column in C,D), where each non-zero entry at position (*i, j*) indicates that LF-MRI feature *i* is matched to CT feature *j* at minimum total cost; (iii) form the element-wise product *C* ⊙ *P* (third column in C,D), retaining only the costs of matched pairs - each non-zero dot’s intensity reflects its individual contribution, and their sum equals the Wasserstein loss. (C, Example 2) The erased CSF regions introduce topological features absent in the CT mask, producing two high intensity bright yellow dots in *C* ⊙ *P* . In the optimal matching plan plot, grey lines connect each matched LF-MRI (green) and CT (blue) feature pair; line length denotes the distance between the pair of points. The red arrows trace the highest-cost matches in *C* ⊙ *P* (the brighter dots) and trace them to their corresponding spatial pairs (grey lines): the matched pairs most responsible for the elevated loss of 0.41. (D, Example 1) The topologically faithful mask yields a low-intensity active dot in *C* ⊙ *P* with a loss of only 0.01, a 4100% reduction relative to Example 2, confirming the sensitivity of the proposed loss to topological inaccuracies even from small structural perturbations.

##### Step 1: Formation of a sublevel Set at a Specific Threshold

The first step in our topological analysis pipeline (Figure 5) involves binarizing the scalar field at a chosen threshold to create a sublevel set. Given a scalar field *S* : Ω → [0, 1] representing the likelihood of CSF in LF-MRI image, we define for each threshold *T* ∈ [0, 1] the sublevel set *B*(*T*) = {*x* ∈ Ω : *S*(*x*) ≤*T*} . This operation naturally creates a binary mask where pixels with scalar values lesser than or equal to *T* are set to 0 (black), while all others are set to 1 (white). The resulting set *B*(*T*) captures regions of high confidence CSF presence at threshold *T* . As shown in Figure 5 (Step 1), different thresholds yield distinct binary masks: at *T* = 0.70, *B*(*T*) preserves a larger region of probable CSF, while at *T* = 0.78, the CSF region shrinks, retaining only the highest-confidence areas. **Step 2: Increasing Collection of Sublevel Sets**. We next construct a *filtration* of the image domain Ω. For the scalar field *S*, we consider the sublevel sets *B*(*T*) for a sequence of increasing thresholds 0 ≤ *T*_1_ ≤ *T*_2_ ≤ · · · ≤ *T*_*n*_ ≤ 1. As *T* increases, these sets form a nested increasing sequence: *B*(*T*_1_) ⊂ *B*(*T*_2_) ⊂ · · · ⊂ *B*(*T*_*n*_). Note that the set *B*(*T*) consists of black pixels, which expands with increase in *T* . This nested structure defines a filtration that encodes the topological evolution of *B*(*T*). As *T* increases, the shape of *B*(*T*) changes: some regions break apart or vanish, and loop-like structures may gradually close or disappear. These changes correspond to the appearance and disappearance of topological features across the filtration. In Figure 5 (Step 2), we illustrate this process using representative thresholds *T* ∈ {0.50, 0.57, 0.64, 0.70, 0.78}, highlighting how topological structures persist or vanish as the filtration progresses. Note that since the range of the scalar field *S* is [0, 1], the threshold also takes values in [0, 1]. **Step 3: Birth and Death of Topological Features**. As we traverse the filtration, we track when topological structures appear (birth) and disappear (death). In particular, we focus on one-dimensional features (*H*_1_), often manifest visually as hole-like structures in the sublevel sets *B*(*T*). Under the adopted visualization, the resulting *H*_1_ features correspond to enclosed CSF cavities or loop-like structures surrounding CSF regions [71], aligning naturally with the clinical interpretation of enclosed CSF cavities. In this setting, these features are interpreted as corresponding to enclosed CSF regions (white areas) surrounded by non-CSF (dark) regions in the sublevel set *B*(*T*). A feature is *born* at a threshold *b* when such an enclosed region first forms in *B*(*b*), and it *dies* at a threshold *d* in *B*(*d*) when the feature disappears. Each persistence pair (*b, d*) lies in [0, 1]^2^ with *d* ≥ *b*, (*b, d*) encodes the lifetime of the feature, and persistence *d* − *b* quantifies feature stability across the filtration. In Figure 5 (Step 3), we show two examples: one feature is present from the beginning (*b* = 0.00) and persists until *d* = 0.65, while another appears later at *b* = 0.62 and disappears at *d* = 0.91.

##### Step 4: Persistence Diagram

The birth–death pairs (*b, d*) for all topological features are collected in a *persistence diagram* (PD). This diagram provides a compact summary of the topological structure of the scalar field. Each point in the PD represents a feature, with its *x*-coordinate indicating the birth threshold and its *y*-coordinate indicating the death threshold. Points near the diagonal (*b* ≈ *d*) correspond to short-lived features that are often associated with noise, while points farther from the diagonal represent more stable structures under the filtration. As illustrated in Figure 5 (Step 4), the PD contains four stable (birth, death) points (0.00, 1.00), (0.62, 0.91), (0.00, 0.65), and (0.57, 0.94), each of which captures the lifetime of a distinct hole-like structure in the white CSF region.

##### Step 5: Topological Loss via Wasserstein Distance

Figure 6 illustrates the importance and step-by-step computation of the proposed topological loss. To enforce topological consistency between the predicted 3D LF-MRI CSF volume 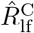 and the ground-truth 3D CT CSF volume 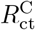, we represent their features using persistence diagrams 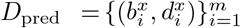 and 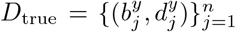 (as explained in the previous steps). As shown in Figure 6(B), the computation proceeds in three steps: (i) we construct a cost matrix *C*, where each entry 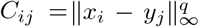 encodes the distance between feature *I* in the LF-MRI persistence diagram and feature *j* in the CT diagram, as visualized in the first column of panels (C,D), with high values (bright dots) indicating strongly mismatched topological features and low values indicating potential spatial correspondences; (ii) we solve for an optimal transport plan 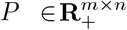 under standard mass conservation constraints [72], where each non-zero entry *P*_*ij*_ (second column in panels (C,D)) indicates that LF-MRI feature *i* is matched to CT feature *j* at minimum total transport cost, thereby defining a soft correspondence between topological structures; and (iii) we compute the element-wise product *C* ⊙ *P* = ∑_*i,j*_ *P*_*ij*_*C*_*ij*_ (third column in panels (C,D)), which retains only the costs of matched pairs, where each non-zero entry reflects the individual contribution of a matched feature pair to the final Wasserstein loss [72], and the sum of all such contributions equals the total transport cost. Intuitively, *C* encodes all possible topological mismatches, *P* encodes how features are optimally paired, and *C* ⊙ *P* = ∑_*i,j*_ *P*_*ij*_*C*_*ij*_ reveals which specific matched pairs actually contribute to the loss. The Wasserstein distance is therefore defined as min_*P*_ ∈Π(*a,b*)*C* ⊙ *P* = min_*P* ∈Π(*a,b*) ∑*i,j*_ *P*_*ij*_ *C*_*ij*_, where Π(*a, b*) denotes the set of admissible transport plans [72]. This measures the minimal “earth-moving” effort required to align the topological signatures between 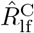 and 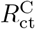. The final topological loss is given by

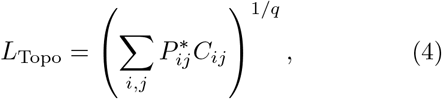

where *P* ^∗^ is the optimal transport plan (diagonal-projected points included in both PDs to balance mass [72]). As illustrated in Figure 6(C), topological corruption (Example 2) introduces spurious CSF cavities absent in CT, leading to high-cost entries in *C* ⊙ *P*, diffuse matching in *P*, and prominent high-intensity contributions (red arrows) corresponding to the largest-error feature pairs responsible for a higher loss (0.41). In contrast, Figure 6(D) shows that a topologically faithful prediction (Example 1) yields near-aligned correspondences, low-intensity entries in *C* ⊙ *P*, and a significantly reduced loss (0.01), demonstrating a 4100% reduction and confirming the sensitivity of the proposed loss to subtle topological perturbations. The resulting loss captures discrepancies in both the existence and persistence of 3D CSF structures (ventricular cavities), and complements pixel-wise supervision by enforcing global anatomical topology in the segmentation. The topological loss is implemented following the formulation in [67], using the *pytorch-topological* library for differentiable computation of persistence diagrams and Wasserstein-based matching.

*An important nuance* in the Wasserstein-based topological loss calculation is that the dominant transport edges in the optimal matching plan do not always correspond to spatially altered regions, though in Example 2 they do. The two removed CSF regions in Example 2 introduce two highly persistent birth-death pairs (two red arrows). Although this is a likely outcome in this particular topology, it is not a general guarantee. The Wasserstein distance minimizes global transport cost; the solver is agnostic to which spatial region generated a birth-death pair. This is further evident in Example 1, where a topologically faithful prediction exhibits a non-trivial transport edge (one red arrow), arising from the global mass redistribution required by the optimal matching, without any anatomical correspondence. While a high topological loss reliably signals topological discrepancy, but the transport plan should not, in general, be interpreted as pixel-level localization of the error.

#### 4) Total Training Loss

The total training loss for EnSegNet-Cross is a weighted sum of four components:

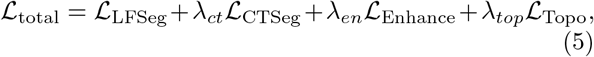

where *λ*_*ct*_, *λ*_*en*_, and *λ*_*top*_ are tunable hyperparameters that control the relative influence of each loss component (see Section Results – Experimental Setup for details). Gradient updates are applied only to *θ*_*E*_, keeping *θ*_*P*_ frozen. This regularized objective enables EnSegNet-Cross to effectively segment LF-MRI with enhancement and topological enrichment from CT.

## Results

### A Experimental Setup

#### 1) Implementation Settings and Training Strategy

We employ five-fold cross-validation [73, 74] for the segmentation tasks. In each fold, 80% of patients were used for model development and the remaining 20% were held out as a test fold for evaluation, with stratified sampling at the patient level to preserve the original class distribution across folds. Within each development split, we further divided the data into training (80%) and validation (20%) subsets for hyperparameter tuning and early stopping. Each scan and its corresponding segmentation mask of size (*H, W*) were transformed into a square image of size (*M, M*), where *M* = max {*H, W*}, to avoid information loss during rotation-based augmentation, which was particularly important due to variability in infant positioning and center shifts. Data augmentation was applied jointly to scans and masks and included horizontal flipping, random in-plane rotations between −20^°^ and 20^°^, cropping, and resizing. Models were optimized using the Adam optimizer [75] with a mini-batch size of 32 for up to 200 epochs, employing early stopping based on validation performance. The initial learning rate was set to 0.001 and decayed by a factor of 0.1 every 50 epochs. Hyperparameters were chosen during the model development phase and set as *λ*_*ct*_ = 1.0, *λ*_*en*_ = 0.005, and *λ*_*top*_ = 0.2, with a weight decay of 10^−6^. All experiments were implemented in PyTorch [76] on an NVIDIA TITAN RTX GPU.

#### 2) Evaluation Metrics

We evaluated performance using two complementary metrics: the Dice Score (Dice) and a novel Volume Score (VScore). Experiments were conducted on the entire dataset as described in Section (Methods A) (denoted as **All**) as well as on a subset of challenging cases with confounding factors (denoted as **Conf**. in the tables). The Dice Score is defined as 1 − Dice Loss, where the Dice Loss has already been introduced in Section (Methods F), capturing spatial overlap between predicted and groundtruth segmentations. For hydrocephalic patients, volumetric accuracy is critical. Prior work [45] reports a mean bias of 2.06% in LF-MRI, reflecting systematic overestimation of ventricular CSF volume, and uses a correlation metric to quantify alignment between CT and LF-MRI volumes. However, these approaches do not explicitly evaluate the consistency of volumetric estimates across modalities. To address this, we introduce the Volume Score (VScore). Let the ground-truth CT volumes be 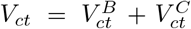, where 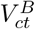 and 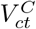 denote brain and CSF volumes, respectively. Similarly, expert-annotated and predicted LF-MRI volumes are 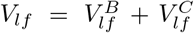, and 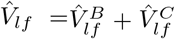, respectively. To compare brain tissue volumes, normalization is required due to differences in total volumes. The normalized predicted brain volume is 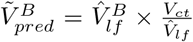, while the normalized annotated LF-MRI brain volume is 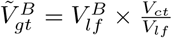. For a dataset of *N* patients, we compute the Pearson correlation coefficients 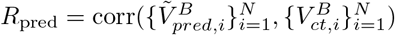 and 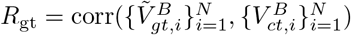, where 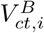 is the CT brain volume of patient *i*. The proposed VScore is

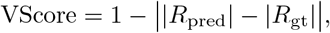

with 0 ≤ VScore ≤ 1, where higher values indicate better alignment between predicted LF-MRI volumes and ground-truth CT volumes after normalization. Absolute values are used in the VScore definition to ensure theoretical boundedness of the metric. Empirically, both *R*_pred_ and *R*_gt_ are consistently positive across all experiments. ||*R*_pred_| − |*R*_gt_|| is used to quantify deviation between the two correlations. Statistical significance of the evaluation (Table 1) was assessed [77], with a significance level of 0.05.

**Table 1.** Comparison of Dice Score (mean ± sd) and VScore (mean ± sd) with the State of the Art Models for LF-MRI Segmentation.

| Model | Dice (All) | Dice (Conf.) | VScore (All) | VScore (Conf.) | p-value |
| --- | --- | --- | --- | --- | --- |
| **EnSegNet-Cross** | $0.8532 \pm 0.03$ | $0.8340 \pm 0.03$ | $0.9318 \pm 0.03$ | $0.9111 \pm 0.05$ | — |
| **EnSegNet** | $0.8003 \pm 0.04$ | $0.7842 \pm 0.07$ | $0.9039 \pm 0.04$ | $0.8894 \pm 0.03$ | $< 0.05$ |
| ResUNet [19] | $0.7751 \pm 0.05$ | $0.6814 \pm 0.06$ | $0.8796 \pm 0.03$ | $0.8701 \pm 0.04$ | $< 0.05$ |
| Swin UNETR [33] | $0.7460 \pm 0.05$ | $0.6688 \pm 0.07$ | $0.8727 \pm 0.03$ | $0.8598 \pm 0.05$ | $< 0.01$ |
| YOLO-SAM 2 [40] | $0.7303 \pm 0.03$ | $0.6943 \pm 0.08$ | $0.8669 \pm 0.07$ | $0.8335 \pm 0.09$ | $< 0.01$ |
| SuperPromptSeg [41] | $0.7269 \pm 0.07$ | $0.6862 \pm 0.04$ | $0.8602 \pm 0.09$ | $0.8271 \pm 0.08$ | $< 0.01$ |

### B. Comparisons against SOTA methods

To rigorously assess the performance of our proposed architectures: EnSegNet and EnSegNet-Cross, we conduct comparative evaluations (Table 1 and Figure 7) against a diverse and chronologically representative suite of state-of-the-art (SOTA) methods in medical image segmentation. The benchmarking cohort spans the evolutionary landscape of medical image segmentation, from early convolutional architectures to contemporary prompt-driven transformer paradigms. **ResUNet** [19] exemplifies early CNN-based frameworks (2012–2019), extending the U-Net architecture with residual connections to improve gradient flow and semantic localization. As the field advanced, **Swin UNETR** [33] emerged during the hybridization era (2019–2021), leveraging a hierarchical Swin (Shifted Window) Transformer encoder integrated with a CNN-based decoder via multiscale skip connections, enabling the fusion of local and global spatial representations. Despite their architectural strengths, both ResUNet and Swin UNETR struggle in LF-MRI due to the inherent ambiguity in anatomical features, particularly for hydrocephalic brains. Convolutional architectures such as ResUNet perform reasonably well in segmenting the CSF region, whose intensity-based and spatially extensive features remain relatively detectable. However, these models often fail to capture the compressed and morphologically distorted brain parenchyma, where neither clear geometric boundaries nor fixed positional priors exist. Transformers, which rely on long-range dependencies and attention to global structure, also underperform due to noise-induced signal loss and lack of strong texture cues in LF-MRI. The advent of foundation models introduced a transformative shift in 2023 with the release of the **Segment Anything Model (SAM)** [37], a segmentation foundation model trained on large-scale prompt-supervised datasets to support zero-shot generalization across imaging domains. Building upon SAM, recent works introduced prompt-generative strategies such as (You Only Look Once) **YOLO-SAM 2** [40], which combines object detection with segmentation by utilizing YOLO-inferred bounding boxes as dynamic prompts for SAM. Complementing this, **SuperPromptSeg** [41] forgoes supervised fine-tuning altogether by employing superpixel-based unsupervised prompting: images are partitioned into semantically coherent regions whose spatial centroids serve as soft supervisory cues to SAM.§

**Figure 7.**
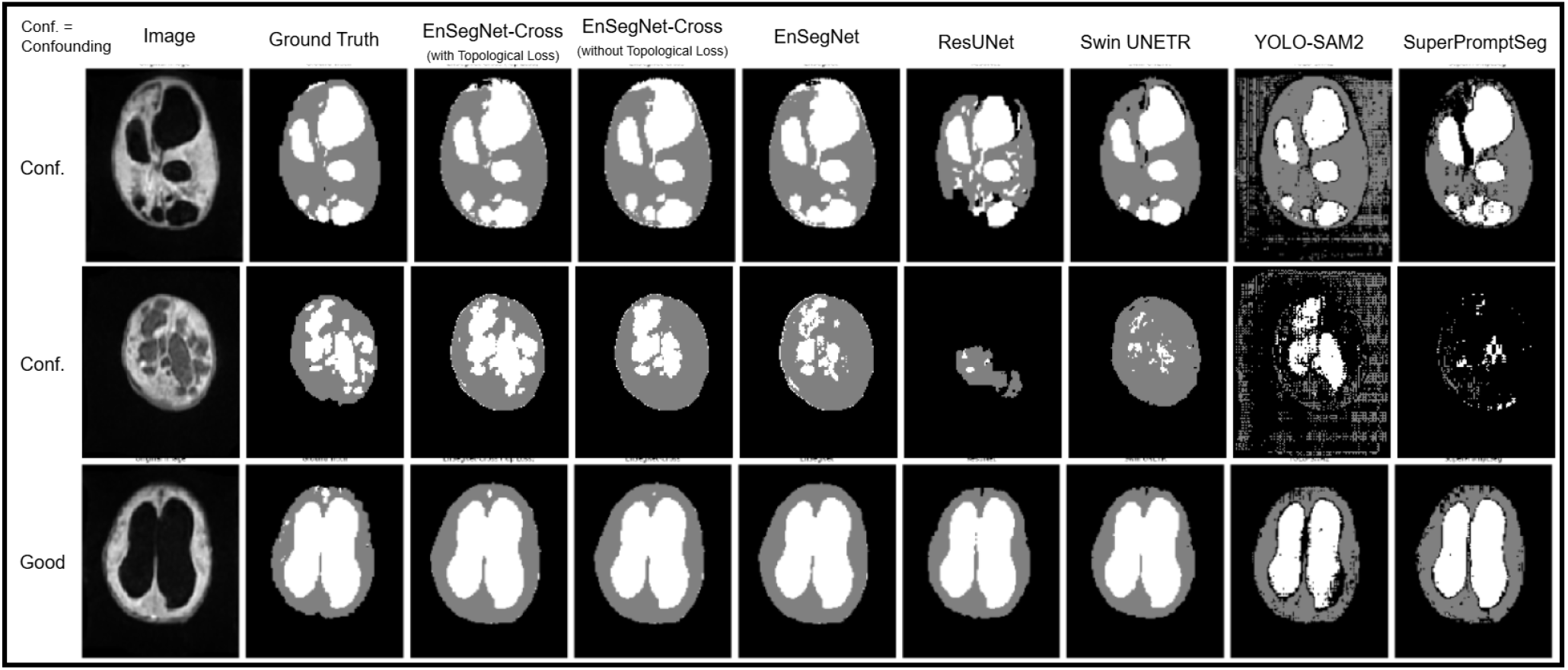
Comparison of the Segmentation prediction with the State of the Art Models with examples in Good and Confounding (Conf.) Cases. Observe that while in the Good case, all the models are performing similarly visually, in the Confounding cases, EngSegNet, EnSegNet-Cross perform much better, leading to better volumetric calculations due to our disease-informed approach.

Table 1 and Figure 7 quantitatively and visually demonstrate that **EnSegNet-Cross** significantly outperforms all competing methods across Dice and Volume metrics (Table 1), particularly in challenging confounding cases (see the second and fourth columns of Table 1, and the first two rows of Figure 7), highlighting the advantage of CT-guided enhancement. Anatomically plausible structures are reconstructed from LF-MRI through cross-modal CT supervision. This effect is further strengthened by topological and statistical regularization, which helps correct distortions that typically mislead both CNN-based [19] and transformer-based models [33]. In contrast, recent SOTA SAM-based methods [40, 41] do not incorporate disease-specific priors into prompt selection. Hydrocephalus induces highly variable anatomical distortions that are difficult to capture using generic prompts. By comparison, both EnSegNet and EnSegNet-Cross explicitly integrate hydrocephalus etiology informed enhancement and prompting strategy, as described in the Methods section. These findings are consistent with [78], which emphasizes the importance of pathology-aware prompting for reliable segmentation using Segment Anything Model in clinically challenging settings.

### C. Ablation Study

To rigorously evaluate the individual and collective contributions of each module in our proposed segmentation pipeline, we perform an ablation study across three configurations: (i) the full **EnSegNet-Cross** model, which integrates both unimodal and crossmodal enhancement, CT-guided supervision (ℒ_*CT Seg*_), and topological loss (ℒ_*T opo*_); (ii) EnSegNet-Cross without the topological regularizer; and (iii) **EnSegNet**, which retains only unimodal enhancement without CT-guided or topological constraints. As shown in Table 2, Dice and Volume scores degrade progressively as modules and regularizers are removed, validating the collaborative importance of enhancement, topology, and cross-modal learning framework. Specifically, the full EnSegNet-Cross achieves the highest Dice (0.8532) and Volume (0.9318) scores, significantly outperforming EnSegNet (0.8003 and 0.9039, respectively), underscoring the critical role of CT-guided learning and structural regularization for segmentation.

**Table 2.** Ablation study of the effect of Cross Module, Enhance Module, and Prompt Module on Dice and Volume Scores.

| Model | $\mathcal{L}_{Topo}$ | $\mathcal{L}_{CTSeg}$ | Dice | VScore |
| --- | --- | --- | --- | --- |
| **EnSegNet-Cross** | Yes | Yes | 0.8532 | 0.9318 |
| EnSegNet-Cross | No | Yes | 0.8314 | 0.9153 |
| EnSegNet | No | No | 0.8003 | 0.9039 |

### D. Interpretability: Explaining the Effectiveness of EnSegNet and EnSegNet-Cross

Importantly, our findings indicate that enhancement is critical not only for improving segmentation accuracy but also for improving the quality and separability of prompts used by SAM. Figure 8(A) shows that distribution matching via the DMM module enables effective CT segmentation, ensuring consistent transfer of CT-based anatomical supervision into the LF-MRI domain. This supervision induces a modality-invariant representation of brain–CSF geometry, allowing EnSegNet-Cross to learn structural priors that are reinforced through cross-modal alignment, leading to more geometrically-consistent predictions compared to LF-MRI-only training in EnSegNet. Figure 8(B) further demonstrates that reductions in enhancement loss strongly correlate with improved segmentation performance and robustness, confirming that enhancement and segmentation are co-optimized in a mutually reinforcing manner. We next show that improved enhancement directly translates into better prompt quality. A compelling evidence is provided in Figure 8(C), which visualizes intensity histograms of superpixel-wise mean intensities (defined in Methods E.2, Prompt Module) for positive (green, brain) and negative (red, CSF) prompt regions: in raw LF-MRI (first row), the positive and negative prompt distributions exhibit substantial closeness in intensity space, making reliable prompt discrimination difficult due to weak separability; with unimodal enhancement (second row, EnSegNet), partial separation emerges, yielding improved segmentation over raw LF-MRI; and with CT-guided crossmodal training (third row, EnSegNet-Cross), this separation becomes most pronounced, with CSF-associated superpixels shifting distinctly toward lower-intensity modes. This progressive distributional separation increases margin in SAM’s prompt embedding space, improving discrimination under degraded imaging conditions. These results are consistent with prior findings on SAM robustness [78], which emphasize prompt conditioning quality. Our analysis extends this by showing that enhancement-induced distributional shifts improve prompt separability, thereby improving segmentation in low-quality LF-MRI. Unlike prior approaches relying on generic prompting, we combine pathology-aware enhancement with CT-derived geometric priors, and UPoLM exploits this improved separability to produce more accurate and robust segmentations.

**Figure 8.**
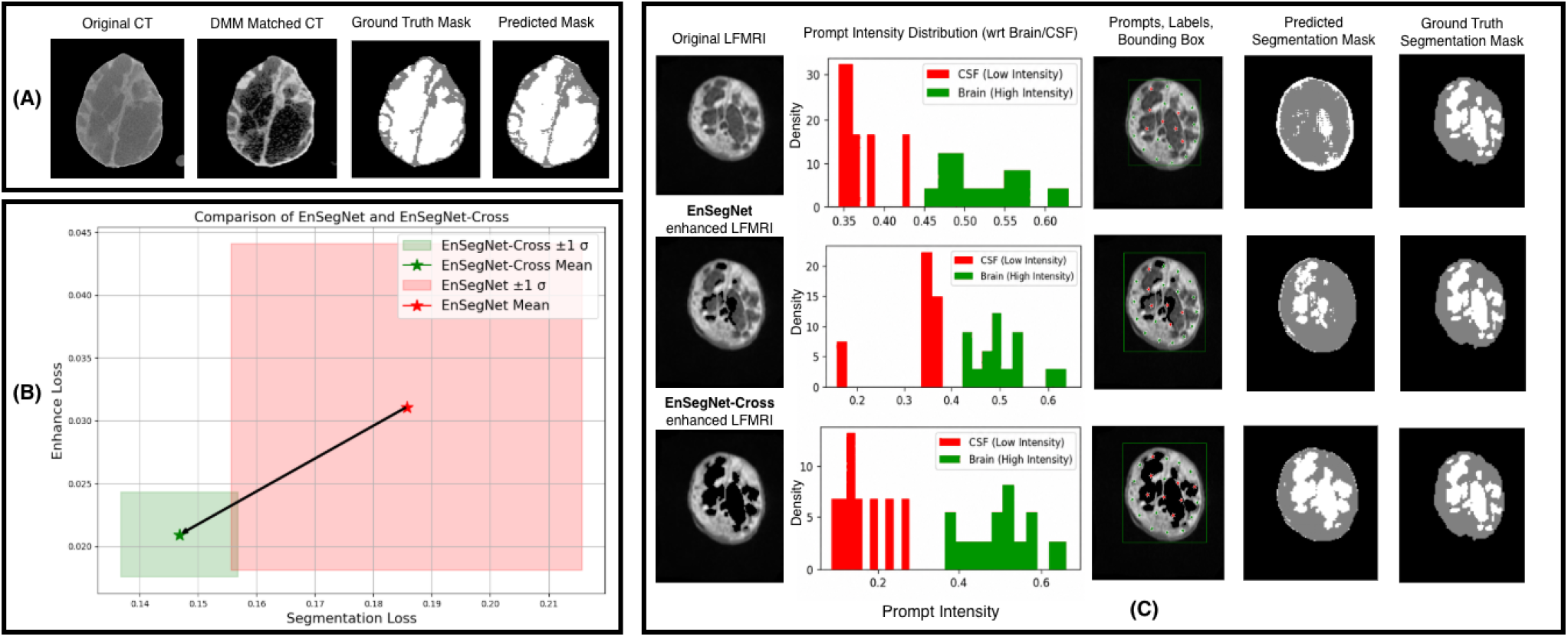
(A) The DMM enables effective CT segmentation producing accurate CT predictions 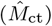 that closely match ground truth. Consistent transfer of CT-based supervision into LF-MRI via DMM induces a modality-invariant representation through cross-modal training yielding more geometrically consistent brain-CSF predictions than LF-MRI-only training in EnSegNet. (B) Reduction in enhancement loss (spurious signal suppression) strongly correlates with better segmentation performance, illustrating co-optimization of enhancement and segmentation. (C) This effect is further reflected in the learned representations, where progressively stronger enhancement leads to clearer separation between CSF and brain superpixel intensity distributions, directly improving prompt separability and downstream segmentation accuracy. Compared to EnSegNet, EnSegNet-Cross exhibits the most pronounced prompt intensity separation, reflecting the additional benefit of CT-guided, pathology-aware enhancement and prompting strategy.

## Discussion, and Conclusion

This study presents EnSegNet-Cross, a topologically enriched framework for accurate and interpretable segmentation for volumetric estimation in hydrocephalic infants with LF-MRI. Our work bridges the fidelity gap between LF-MRI and CT by leveraging CT exclusively during training through a disease-aware Enhancement Module, a topology-regularized Cross Module, and a Prompt Module that provides unsupervised prompting strategy for Segment Anything Model (SAM) to provide high quality segmentation. Results show that EnSegNet-Cross consistently outperforms SOTA methods like CNN [19], Transformer [33], and prompt-based SAM baselines [40, 41], across Dice and Volume metrics, most pronouncedly in challenging cases.

### Limitations and Future Works

1) A key challenge is that raw Hyperfine acquisitions are inaccessible: a proprietary enhancement algorithm, subject to change across software versions, transforms the raw signal into the scans on which all downstream processing operates. EnSegNet-Cross is robust under a mild regularity condition: if the Hyperfine enhancement map is homeomorphic, it preserves anatomical topology, inducing only continuous deformations of true structures. Our cross-modal topological loss implicitly corrects any residual discrepancies by aligning predicted LF-MRI topology with CT ground truth. Critically, this correction operates directly on the already-enhanced scans: because our training aligns predicted LF-MRI topology with unenhanced CT ground truth, any smooth and continuous transformation applied upstream by the Hyperfine pipeline is implicitly absorbed and corrected, even without access to the raw signal. This provides a concrete advantage over pixel-wise approaches, which have no mechanism to recover upstream global structural errors. Notably, this smoothness assumption is not merely a theoretical convenience. Prior works [79, 80] employing deep learning reconstruction pipelines to ultra-low field images, that are by construction smooth differentiable mappings, imply that the upstream transformations they induce are continuous deformations of the underlying anatomy and thus do not perturb its topology. Limits exist, however: if the proprietary map is not homeomorphic, e.g., it ‘cuts or pastes’ intensity regions, topological correction will fail. 2) Orientation-dependent topological variability presents a second challenge: because the head is a rigid body that cannot be identically repositioned across repeat scans or across machines, thin CSF connections may appear or disappear depending on scan angle, introducing spurious gradients into the topological loss, illustrated in Figure 6, where removing two small connected regions increases topological loss by many orders of magnitude. The principled remedy is registration to a common reference frame prior to topological loss computation. Recent work [81] demonstrates that geometric distortion, rather than resolution, is the primary source of registration failure in real Hyperfine data, and that a synthesis-based pipeline achieves the most robust performance across scanner configurations, making it a natural upstream complement for future work. 3) An interesting open question is whether cross-modal training can further improve the Hyperfine proprietary enhancement itself. Our method implicitly corrects enhancement induced topological artifacts under the smoothness condition above; the deeper question is whether, given access to raw pre-enhancement acquisitions paired with CT, the Enhancement Module could learn principled intensity mappings toward anatomically grounded targets and perform better enhancement. A principled path toward this is explicit noise modeling: LF-MRI noise is well-characterized as a Rician distribution [82, 83], and incorporating this statistical prior directly into the Enhancement Module would provide a physically grounded basis for intensity mapping, enabling the network to first estimate and separate the spatially varying noise floor from true anatomical signal before applying CT-guided cross-modal correction, yielding enhancement targets that are both noise-calibrated with LF-MRI and structurally consistent with CT-derived anatomy. This twostage approach, noise-aware signal recovery followed by CT-guided topological alignment, is a natural extension of our cross-modal framework. Current limitations include dataset scale (47 subjects, single site), absence of explicit geometric registration, opaque and mutable scanner preprocessing, and 2D slice-stacking rather than true volumetric processing. These limitations define concrete directions for future work. Our work EnSegNet-Cross is a substantial advance demonstrating that accurate, high-fidelity, radiation-free volumetric monitoring is achievable from LF-MRI.

## Acknowledgements and Conflict of Interest

This work was supported by NIH grant 5R01HD085853-12 (ClinicalTrials.gov registration number NCT01936272), and an Equipment Transfer Agreement between Hyperfine, Inc and Yale University. The authors declare no conflicts of interest.

## Data Availibility

The data cannot be made publicly available because they contain sensitive personal information. Limited data that supports the findings of this study are available upon reasonable request to the authors.

## Footnotes

‡ The dataset for this study was collected at CURE Children’s Hospital of Uganda (CCHU), a nationwide neurosurgical referral center located in Mbale, in the eastern region of Uganda.

§ Note that our SOTA comparisons do not include domain adaptation approaches [55] because those approaches do not have associated target domain ground-truth labels for network training, i.e, LF-MRI ground-truth segmentation, making it a distinct problem statement from ours.

